# Informing University Covid-19 Decisions Using Simple Compartmental Models

**DOI:** 10.1101/2021.07.01.21259851

**Authors:** Benjamin Hurt, Aniruddha Adiga, Madhav Marathe, Christopher L. Barrett

## Abstract

Tracking the COVID-19 pandemic has been a major challenge for policy makers. Although, several efforts are ongoing for accurate forecasting of cases, deaths, and hospitalization at various resolutions, few have been attempted for college campuses despite their potential to become COVID-19 hot-spots. In this paper, we present a real-time effort towards weekly forecasting of campus-level cases during the fall semester for four universities in Virginia, United States. We discuss the challenges related to data curation. A causal model is employed for forecasting with one free time-varying parameter, calibrated against case data. The model is then run forward in time to obtain multiple forecasts. We retrospectively evaluate the performance and, while forecast quality suffers during the campus reopening phase, the model makes reasonable forecasts as the fall semester progresses. We provide sensitivity analysis for the several model parameters. In addition, the forecasts are provided weekly to various state and local agencies.

## 1 INTRODUCTION

The COVID-19 pandemic has presented an extreme challenge for the global human population. With a high mortality rate and significant economic impact, the drive to learn from and develop techniques to assuage the damage of COVID-19 is at the forefront. While the initial phase of the pandemic was marred by lack of understanding of the dynamics of the disease spread and a lack of coordination, subsequent efforts to mitigate the spread have been marked by informed policy-making. Some of the key efforts have been in forecasting of cases, deaths, hospitalization, etc. and has involved top academic institutions and modeling teams (ForecastHub 2020).

In order to reduce inconsistency of forecasts between teams, these forecasting efforts enforce standardized data sets, forecast formats, and evaluation metrics. However, these efforts have largely focused on national-, state- and county-level forecasting. This work notwithstanding, there is an outstanding need to understand the course of the pandemic among a variety of other resolutions. To this point, since the start of the epidemic, there have been over 530,000 cases reported by American colleges and universities (Times 2020). As a result of rising case counts, universities have had to move much of their teaching to online modalities given that campuses are subject to high contact rates among individuals. However, some of the activities within universities require physical presence and hence ensuring the safety of the staff and students is a major challenge.

In this paper, we describe our efforts modeling and forecasting cases on multiple university campuses depicted in Figure 1. To the the best of our knowledge, this is one of the only campus-level real-time forecasting efforts at present. Beginning with the Fall 2020 semester, we have been providing continuous up-to-date campus-level weekly COVID-19 cases forecasts and scenario analyses of four Virginia universities, shown in Table 1, to state and local officials in order to inform policy relating to pandemic mitigation (e.g. group sizes, class openings, service availability, student movement) and student testing frequency. By providing multiple scenarios, our decision makers received a view of not only the course of the pandemic in real-time, but also the upside and downside potential cases from decreases and increases in the student contact rate, thus informing contingency planning.

**Table 1:** The enrollment of the four modeled schools along with the population of the county of city where they are located.

| School | Enrollment<br>(2018/2019) | County/City | County Population<br>(2019) |
| --- | --- | --- | --- |
| Virginia Tech (VT) | 34,683 | Montgomery | 98,535 |
| Virginia Commonwealth University (VCU) | 30,697 | Richmond | 226,622 |
| University of Virginia (UVA) | 24,639 | Charlottesville | 47,096 |
| James Madison University (JMU) | 21,751 | Harrisonburg | 53,273 |

**Figure 1:**
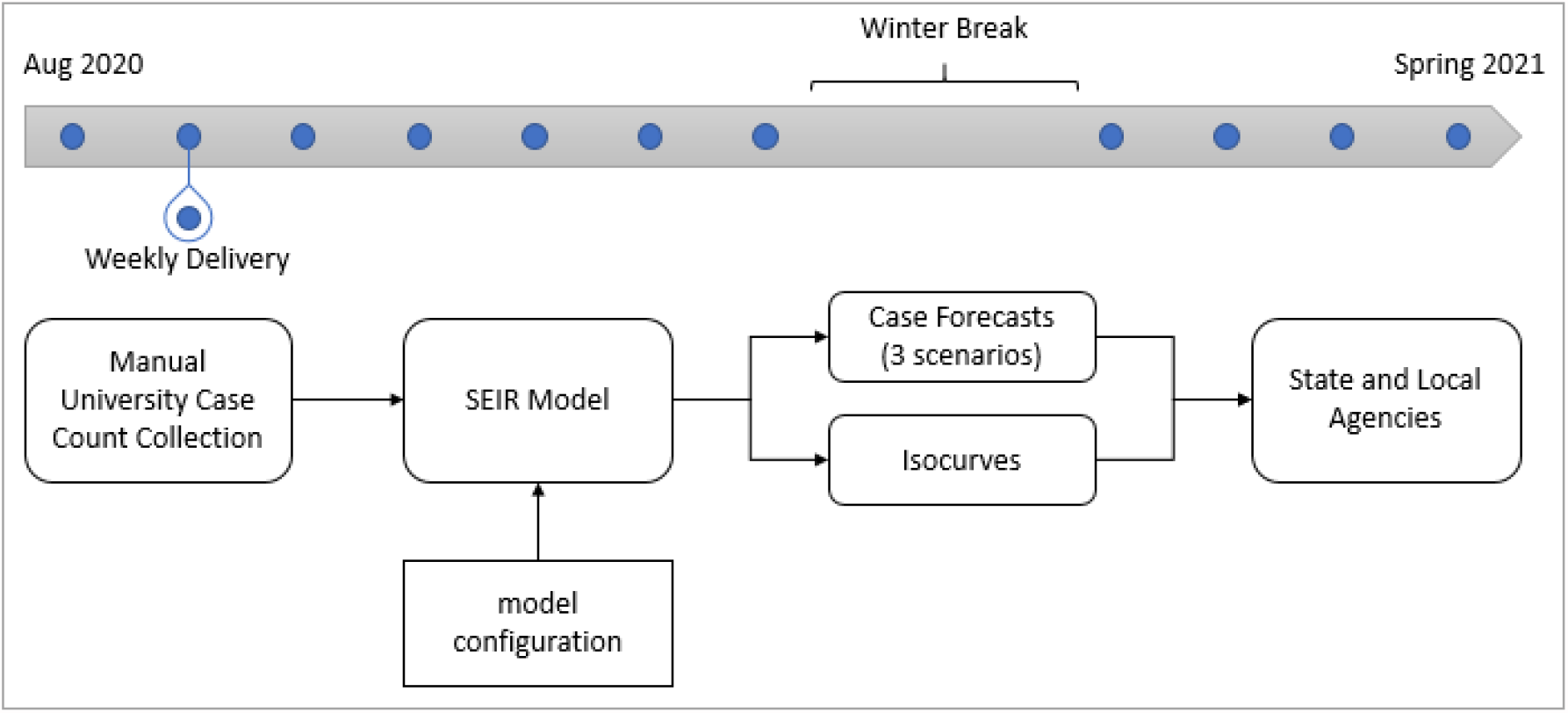
The weekly work flow.

During our work, we faced a variety of data issues, including: collection, inconsistency of reporting frequency, formats, and resolution, restatements, and corrections. Many of these data issues resulted from the difficult positions that colleges found themselves in. Building a team to address the pandemic, developing new reporting systems, testing individuals, and disseminating that information to their community all required time and resources that were unlikely to be budgeted for. Given the lack of and challenged nature of the campus case data, statistical or purely data-driven models were not feasible at the time. As a result, we employed the Susceptible-Exposed-Infected-Recovered (SEIR) model, a popular infectious disease compartmental model which incorporates the causal mechanisms of disease spread dynamics. We employed one SEIR model per school with the model parameters calibrated on the respective school’s case time series. The model produced two week’s worth of daily forecasts and for performance evaluation purposes we aggregated them into two weekly forecasts.

Despite these challenges, our weekly report contained: 2 weekly COVID-19 case forecasts under each of three scenarios (stable contact rate, contact rate halved, and contact rate doubled), along with isocurves, described in (Weitz, Joshua 2020), that informed intervention efforts by depicting the trade-off between testing frequency and contact rate in resulting new cases. The case forecasts and testing intervention scenarios are combined to create an important and valuable source of information for COVID-19 management decision makers.

### 1.1 Related Work

SEIR models have a long history. They have been a primary candidate for COVID-19 modeling (COVID-19 ForecastHub 2020), and, while they have performed generally well, there have been some efforts to use either statistical or other mechanistic models alongside them for pandemic research (Adiga et al. 2020; Yang et al. 2020; Xiao et al. 2020). Work involving their use for COVID-19 forecasting is already widespread (He et al. 2020), including those studying the difficulties that SEIR models have making such forecasts (Roda et al. 2020). Despite the difficulties, SEIR models have, by now, been used to forecast COVID-19 cases all over the world in both large- and small-scale resolutions (Annas et al. 2020; Radulescu et al. 2020; López and Rodo 2021). These studies also, together, investigate the effect of various intervention measures (closures, vaccinations, isolation, etc.). Additional intervention studies include the placement of recovered individuals in high-contact roles (Weitz et al. 2020), and optimizing isolation control policies based on improved testing (Li et al. 2020).

Along with the work done at higher resolutions on forecasting and intervention techniques, there have been several investigations concerning college campuses and COVID-19. Due to to the lack of data and understanding of university reopening strategies and testing protocols, initial work focused on understanding effects of interventions using hypothetical cohorts (Paltiel et al. 2020) to understand the effects of pre-arrival testing, regular testing, and test specificity, etc. Other works described the progression and spread of the virus through a college campus (Wilson et al. 2020). Further works focused on studying college administration detection and mitigation techniques and the impact on observed cases (Fox et al. 2021; Betancourt et al. 2021; Marsicano et al. 2020; Walke et al. 2020; Losina et al. 2021). There has been work completed that delves into the impact of such actions by schools on the health of the student population itself, often focusing on mental health (Conrad et al. 2021; Zhai and Du 2020). Additionally, there has been work completed on the impact of large schools on the case prevalence of their surrounding community (Leidner et al. 2021; Mehrab et al. 2020; Andersen et al. 2020), finding that schools with students returning to campus may have contributed to higher observed cases in the broader community. These findings heighten the importance for systems, like the one described in this article, that can inform college administration-level decision making.

The paper is organized as follows: In Section 2 we discuss the challenges of data curation, model details, scenarios, and probabilistic forecasting. In Section 3, we present the forecasts, discuss intervention modeling, evaluate forecast performance using multiple metrics and perform sensitivity analysis of the SEIR model parameters. We conclude the paper with a discussion in Section 4.

## 2 COLLEGE CASE FORECASTING

### 2.1 Data Collection

To identify the initial subset of US schools to model, we used a combination of a list of schools made available by the New York Times (Times 2020) with the most cases, along with the largest schools in each state. Given the lack of a central case database at the time, we decided to collect the data manually from each school’s COVID-19 dashboard.

Throughout the modeling period, collecting data reliably posed a challenge. At the outset, not all of our chosen universities made their COVID-19 case data publicly available and we were simply unable to acquire case data from these schools regularly. Given the number of schools we were able to find data for, eventually 67 institutions, we decided to not pursue extra efforts to track cases from additional schools. When we set up the pipeline to provide forecasts to various agencies in the state of Virginia, we settled on modeling just four of the largest public schools in Virginia.

Once the subset of schools, with publicly available COVID data, was determined we were next confronted with the reality that there was no consistent periodicity, format, or granularity between the schools that made their COVID cases available. Case reporting frequency could be daily, partial weekly, weekly, or irregular. The format varied from static images of case counts to Tableau windows, from simple data tables to interactive charts. The granularity of reporting confirmed cases varied from a single number for the school, to subsections shown for each of students (on- and off-campus), faculty, and affiliates. As a result of all these variations, we manually collected data from each school’s COVID-19 dashboard. Scraping techniques were investigated, however, given the individual nature of each reporting method, this was not a scalable solution. These issues, shared among those modeling college-level cases, have led to initiatives to centralize college data (The College Crisis Initiative 2020).

Beyond the variations between school postings of COVID-19 cases, we also encountered the issue of frequent restating of a school’s case history. Usually minor updates occur to the latest posted data as cases are confirmed late, or lag the initial release of that day’s data. Occasionally, though, restatements several days or, rarely, even weeks in the past would occur. In these cases we updated the school data to the latest. To dampen the effects of variability in daily reporting and time biases, we applied a rolling 7-day average to the daily case time series.

The challenge that coping with the pandemic posed to schools explains some of the data collection difficulties. The schools had to build teams to inform the plethora of decisions to be made, build infrastructure for collecting cases, monitor a decentralized community of on- and off-campus students with varying levels of commitment or interest in following pandemic policies, and create new reporting systems or dashboards to make that information available. Given a limited set of resources, none of these are easily done on short notice especially in an environment when the management of the school itself is under a variety of new pressures (budgetary, when/if to open for in person classes, what to do with on-campus students, etc.).

### 2.2 SEIR Model

We employ a compartmental Susceptible-Exposed-Infectious-Recovered (SEIR, shown in Figure 2) model with outcome processing for case confirmation generated at the school-level resolution. The campus population is divided into the four compartments and the rate at which the individual compartment population changes are given by

**Figure 2:**
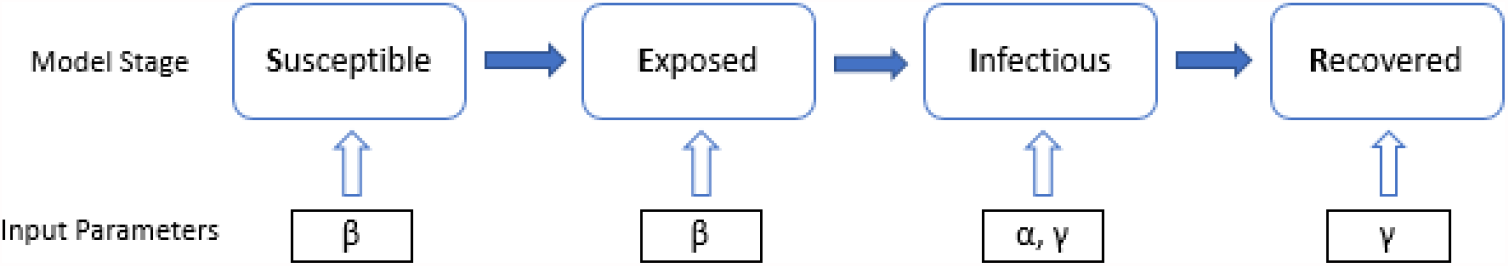
The SEIR model and the location of the input parameters.

**Figure 3:**
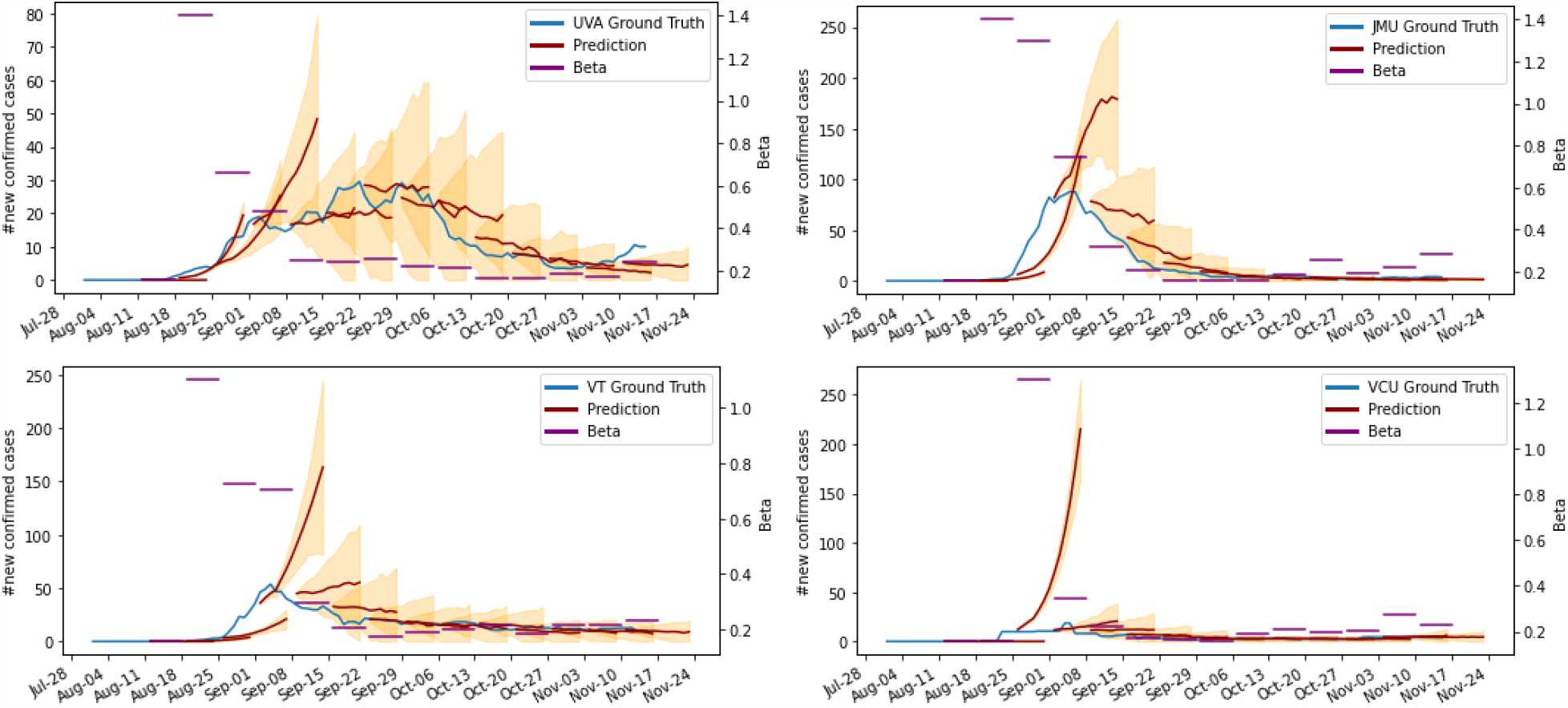
The figure shows the 7-day rolling average daily new COVID-19 cases (blue line) during fall semester (August through mid-November) of the four different Virginia universities. Also shown are the weekly forecasts from the SEIR model (dark red lines). Each forecast is made two weeks forward. The orange shaded area represents the 95% confidence interval for each prediction. Finally, the purple lines represent the future beta used by the SEIR model for each predictions.

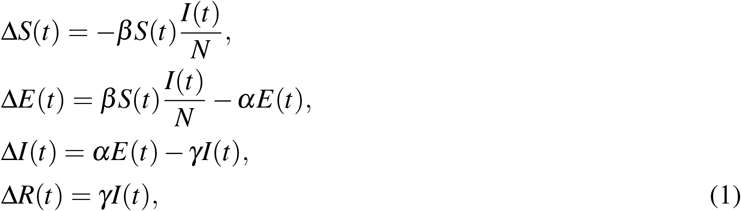

where *β, α*, and *γ* are the transmissibility, incubation rate, and recovery rate parameters, respectively, and indicate the disease specific parameters. The model employed in this paper is an adaption of the non-network version of PatchSim model (Venkatramanan et al. 2019). In this model, we train each school in isolation where the interactions, effects of social distancing and miscellaneous adaptations are captured as temporal variations (daily variations) in the transmissibility term *β* while keeping other parameters, the duration of incubation (5 days) and recovery rate (5 days), case ascertainment rate (7x, each confirmed case indicates 7 underlying infections in the population), and delay from exposure to confirmation (7 days) fixed (CDC 2020). Widespread pandemic eliminates sensitivity to initial conditions, hence we assumed steady low-level of importation/external seeding (1 case per 10 million).

Let the SEIR model be expressed as Δ**S**,Δ**E**,Δ**I**,Δ**R** = *f*_*α,γ*_ (***β***), where boldface indicates the vector form of a time series, for example, Δ**S** = [Δ*S*(0), Δ*S*(1), …, Δ*S*(*T*)] and Δ*S*(*i*) = *S*(*i*) *− S*(*i −* 1) indicates the change in number of susceptibles from day *i −* 1 to *i* and ***β*** = [*β* (0), *β* (1), …, *β* (*T*)]. Note that, in practice, Δ**S**,Δ**E**,Δ**I**,Δ**R** are typically not observed but only the reported number of cases **C** is available. We typically employ the case ascertainment value *κ* that is the average number of infections getting reported, that is, **C** = *κ***I**. For the sake of brevity, we denote the SEIR model as Δ**C** = *f*_*α,γ,κ*_ (***β***). With the assumed disease parameter values the transmissibility parameter is calibrated as

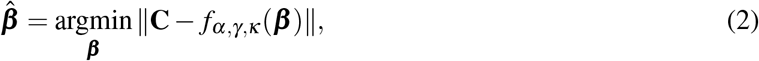

where the nonlinear optimization problem is solved using Broyden-Fletcher-Goldfarb-Shanno algorithm, a gradient-based optimization of the mean-squared error objective function. This method allows for layering counterfactual projections (i.e., increase or decrease in future *β*). Once calibrated, the forecasts are obtained by projecting forward the 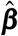 in multiple ways and is discussed in Section 2.3.

**Testing** Most universities resorted to high volumes of testing during the fall semester. The tested individuals identified as infected are subjected to isolation which effectively results in the reduction of number of infected individuals in the population. Key to testing is the frequency at which they are conducted and also the test sensitivity (true positive rate). Given the test parameters, the effects of testing on the disease dynamics can be incorporated into the SEIR model as follows:

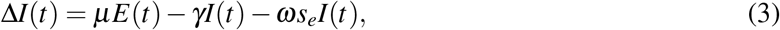

where *ω* is the rate at which an individual is tested and *s*_*e*_ is the sensitivity of the test (was determined to be 75%). In our modeling efforts, we incorporated a range of testing rates and generated possible scenarios that are captured in isocurves, shown in Figure 4.

**Figure 4:**
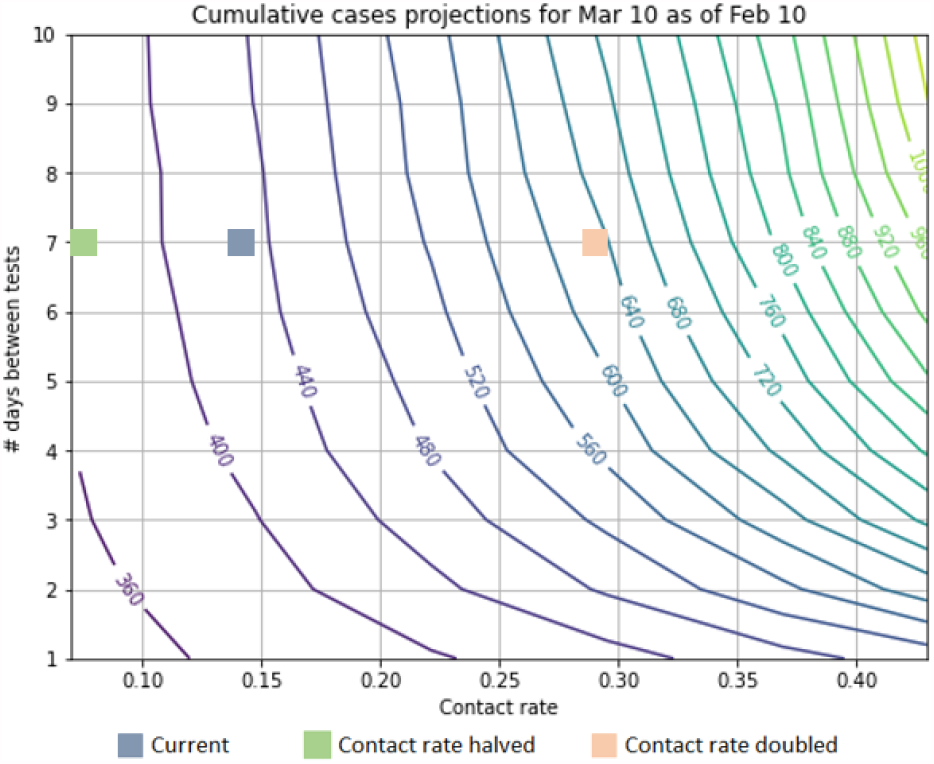
The isocurves show the resulting cases four weeks forward (curved lines) given the frequency of testing (y-axis) and contact rate (x-axis). As testing frequency increases, a higher contact rate can be tolerated to result in the same number of cases (due to the resulting mitigation measures taken for persons testing positive). Alternatively, if the contact rate decreases, the school can choose to decrease the rate of testing in order to maintain an expected new case count that it is comfortable with. The three colored squares indicate the position of the school’s current testing policy and contact rate.

### 2.3 Scenarios

Beyond the data collection issues, predicting 1- and 2-weeks forward cases for a college campus is difficult due to the highly interactive structure of campus life. In order to account for the potential for rapid changes in contact rate we investigated avenues that inform forward contact rate adjustments.

One of these avenues was to use location-based mobility data. Studies of this have shown promising results for the early period (reopening) of a semester (Mehrab et al. 2020), but the correlations did not hold for later portions of the semester.

Another avenue was to use knowledge of upcoming events on campus (e.g. Spring Break, Greek-life rush) that we believed would result in higher contact rates among the students to alter the forward beta. We found it too difficult to reliably model these events accurately ahead of time - simply having awareness of such events along with understanding their size, and the interaction dynamics was not feasible within the time constraints of our process.

Given these difficulties, we produce three different scenarios: after calibrating the model on the known case data, we compute the average of the latest 7-days of ***β*** and use that to produce the forecasts for Scenario 1.0, that is 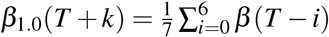, for *k* = 1, 2, …, 14. The two other scenarios are: Scenario 0.5 - the “best” case where we reduce Scenario 1.0’s future *β*_1.0_ by half, representing some combination of the success of containment policies and student compliance. Scenario 2.0 - the “worst” case where we double Scenario 1.0’s future *β*_1.0_, representing the potential contact rate spike resulting from a particular social event or perhaps a breakdown of policy compliance in general (e.g. due to a change in weather, COVID fatigue, etc.).

### 2.4 Probabilistic forecasts

Of high importance in case forecasting is the need for quantifying the uncertainty. Unlike the point forecasts, probabilistic forecasts provide a model’s confidence in its forecast values. We determine uncertainty in our model by perturbing the projected 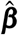 values as a function of the training error (calibration error). Specifically, we randomly sample *N* times from a Gaussian distribution with mean as calibrated *β* and variance as the training mean square error and project forward to obtain *N* different forecasts. We then compute the statistics of the *N* forecasts.

We also compare the SEIR model to an ARIMA model, which does reasonably well for time series forecasting. Given the short time period and continual nature of the prediction, we built an individual non-seasonal model for each of the forecast periods and school time series. Each model uses a training period length of 15 days of each school’s log-transformed daily new COVID-19 cases time series. The model’s p, d, and q parameters are tuned by optimizing the Bayesian Information Criterion over a range of [0, 10] for p and q, and [0, 2] for d. The prediction accuracy can be found in Figure 5. It can be seen that the ARIMA model performs fairly in-line with the SEIR scenarios 0.5, and 1.0. The reason for employing the SEIR model as opposed to the ARIMA model, is the ability to incorporate multiple scenarios based on the same input data. For decision makers faced with planning contingencies having a model that can give reasonable bounds to a “worse” scenario is vital to forward planning.

**Figure 5:**
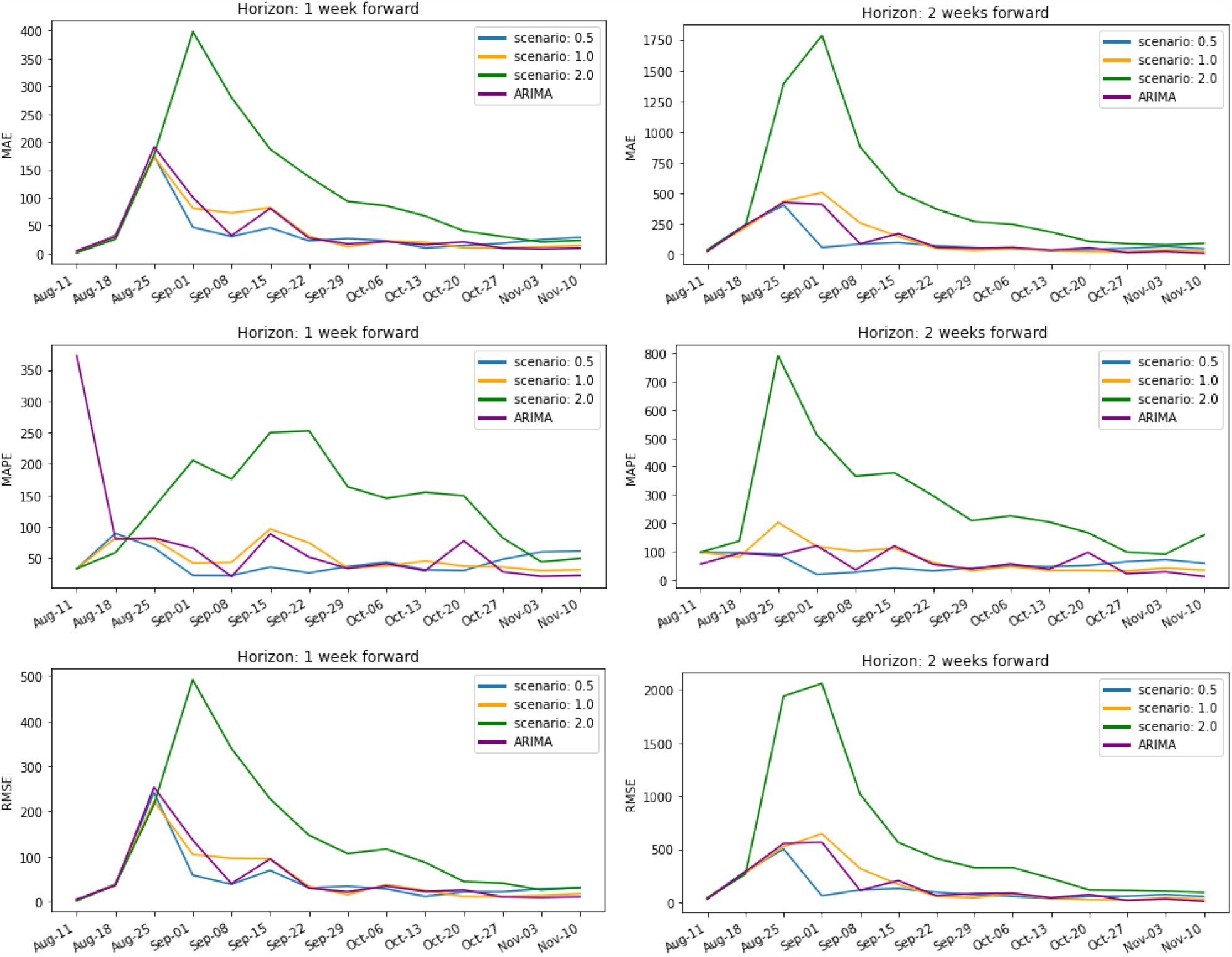
The above shows three different mean error measures, MAE, MAPE, and RMSE, for predictions from the four Virginia colleges for the 1-week and 2-weeks forward predictions. The different lines are the different contact rate scenarios run for each school: SEIR constant, halved, doubled, along with the ARIMA model for comparison. The MAPE calculation is corrected by adding 1 to the denominator in order to avoid division by 0. The initial spike in the errors can be observed in all scenarios and ARIMA and across both forecast periods and measures. Afterwards, MAE and RMSE drop as the magnitude of cases dwindles.

## 3 MODEL PERFORMANCE

We present forecasts for four Virginia universities in Figure 3. From the figure, it can be observed that in the initial weeks most universities show sharp increase in cases. The sharp rise coupled with lack of sufficient historical data makes it hard for the model to train and hence does not closely predict the course of the pandemic at the school. Once the initial phase of high weekly cases passes, the model forecasts appear to closely capture the trajectory across all campuses. We quantify the performance of the model using Mean Absolute Error (MAE), Mean Absolute Percentage Error (MAPE), and Root Mean Squared Error (RMSE). The forecast performance across the fall semester based on these metrics is presented in Figure 5 (Note that for evaluation purposes we aggregate the daily forecasts to weekly forecasts which helps overcome the reporting biases observed over different days of the week). The MAE and RMSE show error reducing after the initial period for both one-week- and two-week-ahead forecast horizon. The errors corresponding to Scenario 2.0 are consistently higher than the other two scenarios thus indicating that the cases were mostly below the “worst case” expectations. As for Scenarios 0.5 and 1.0, the MAPE shows steady performance of the models indicating that the errors were generally proportionate to the cases. We also compute the number of times the ground truth falls within the 95% confidence interval of the forecasts. Apart from JMU, the 1.0 scenario was able to capture the trend in one-week-ahead cases nearly 50% of the time while the one-week-ahead case trend was captured 60-90% of the time by at least one of the scenarios. This trend holds, although at higher percentages, for the two-week-ahead prediction coverage. Given that all three scenarios jointly have a higher percentage of ground truth coverage demonstrates the value of multiple scenarios.

As a result of the weekly delivery schedule, we have a relatively short amount of time to produce our results and make changes to the model. The consistency of this schedule was an important tool for the recipients to monitor the course of the pandemic in their purview. In addition to the short time frame, the schools’ COVID-19 policy was dynamically updated throughout the period. These updates (e.g. to the rate of testing or various mitigation practices) resulted in the need to constantly change the model to be consistent with the current state of the college’s efforts.

### 3.1 Sensitivity Analysis

The results presented in Section 3 are based on fixed SEIR model parameters discussed in Section 2.2. The *β* (*t*) was calibrated based on the model parameters such as incubation period, recovery period and delay from exposure to confirmation. A natural question that arises in this context is the variability of a model’s forecast performance based on the choice of the parameters. In order to determine the sensitivity to the choice of parameters, we construct a full factorial design experiment using a range of parameter values: Incubation period ({3, 5, 7, 9} days) and recovery ([3 *−* 14] days in steps of 1 day). The model is calibrated over every combination of these parameters and then run forward for 14-days for the three different scenarios (0.5, 1.0, and 2.0). This experiment is repeated for each submission week and the corresponding MAE is determined. We present the distribution of the MAE obtained for a fixed value of incubation period and recovery period using boxplots (depicting the quartile points) in Figures 6 and 7, respectively. Each boxplot represents a group of model forecasts that employs the indicated model parameter in the x-axis. Visual inspection indicates low variability in performance across groups for Scenario 1.0 and 0.5. However, for Scenario 2.0 the variation in model performance is quite significant. In order to determine if there is statistically significant differences between the groups across all scenarios, we perform a one-way analysis of variance (ANOVA) test. The one-way ANOVA test compares the means between the groups and statistically determines whether the means are significantly different from each other. We present the *p-values* of the test in table 2. A critical value of 0.05 was chosen and hence *p >* 0.05 indicates the null hypothesis that the samples in all groups are drawn from populations with the same mean values. The entries in the table indicate that for Scenarios 0.5 and 1.0 the variability in model performance for various design choices is statistically insignificant. Hence, the choice of incubation period of 5 days and a recovery period of 5 days is a reasonable assumption.

**Table 2:** ANOVA test *p-values* to determine significance of the variation of the mean performance across parameters. The columns indicate the scenarios while the rows indicate the parameter with respect to which SA is performed. For a given parameter and scenario of interest we obtain a group of absolute error values. If *p >* 0.05 then the null-hypothesis, that the means of different groups are statistically equal, is true.

|  | 1.0 | 0.5 | 2.0 |
| --- | --- | --- | --- |
| Recovery period | 0.63969 | 0.999999 | 1.02654e-06 |
| Incubation period | 0.995204 | 0.994067 | 1.46495e-12 |

**Figure 6:**
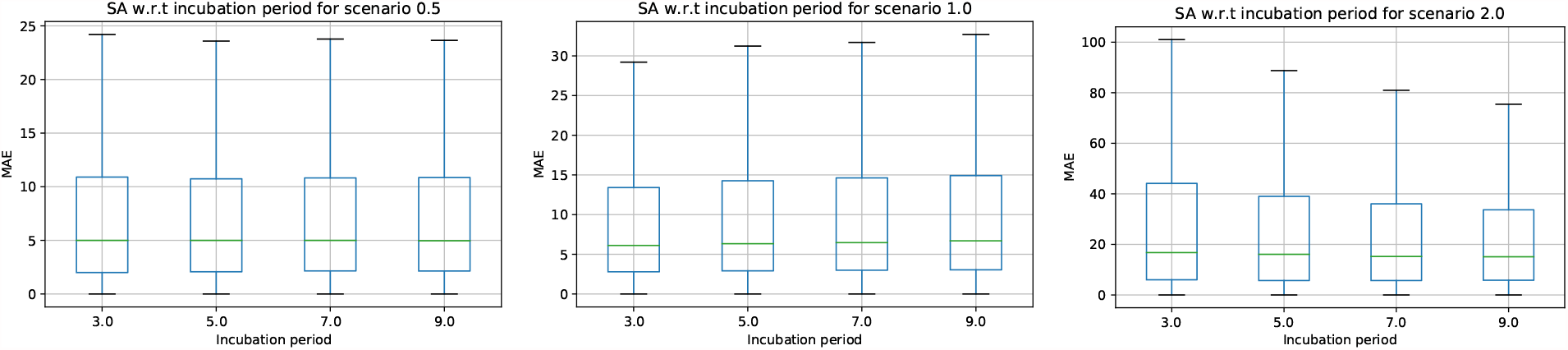
Sensitivity analysis to determine the effect of changing the incubation parameter. The boxplots indicate the variation in MAE for different incubation periods across the three scenarios.

**Figure 7:**
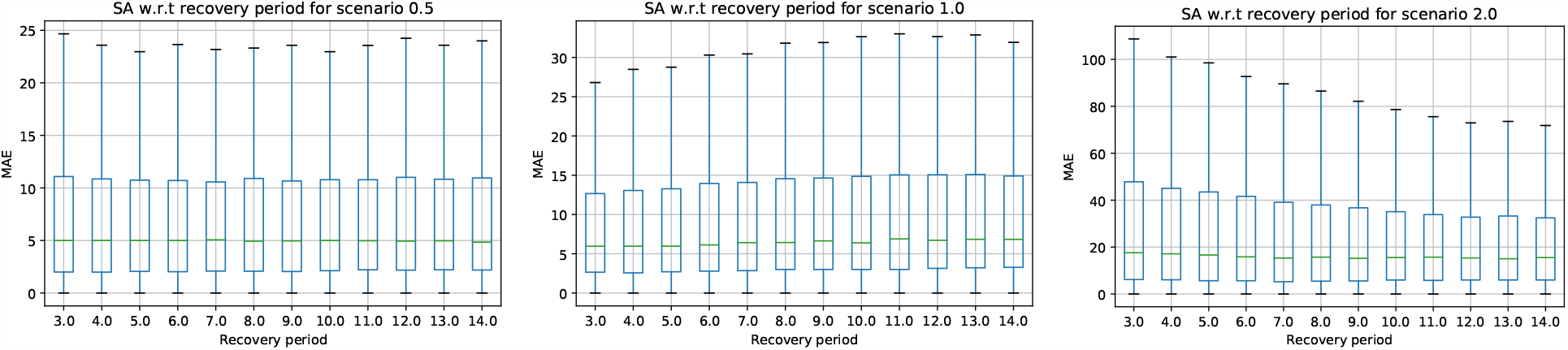
Sensitivity analysis to determine the effect of changing the recovery period parameter. The boxplots indicate the variation in MAE for different recovery periods across the three scenarios.

## 4 DISCUSSION

In this paper we present a real-time campus-level forecasting framework. In terms of the performance, the initial spike in error results primarily from two factors: a training period that is too short - the model has no to very little case history - and the first phase of the epidemic on campus being the period when students are gathering from widespread locations (e.g. UVA students came from 44 US states, and over 40 countries (Collegefactual 2021)). This first phase results in the most weekly cases observed and a rapidly changing slope of the new-case curve.

During the period after the model has been trained on a reasonable amount of data, the forecasts provide, at a minimum, a directionally correct indication of the trajectory of cases for the universities, shown in figure 8. All three scenarios jointly have a higher percentage of ground truth coverage which demonstrates the value of having multiple scenarios. The ground truth coverage of the scenarios together is important since this process is implemented on a real-time basis and used for decisions being made week-to-week. The decision makers consuming these forecasts are able to have a reliable view of the forward expected cases, upside, and downside potential of the new cases in the future which allows them to alter the measures that influence the tolerable future contact rate (e.g. managing group gathering sizes, in-person class and program availability, dining hall arrangements, and mobility from and around dormitories).

**Figure 8:**
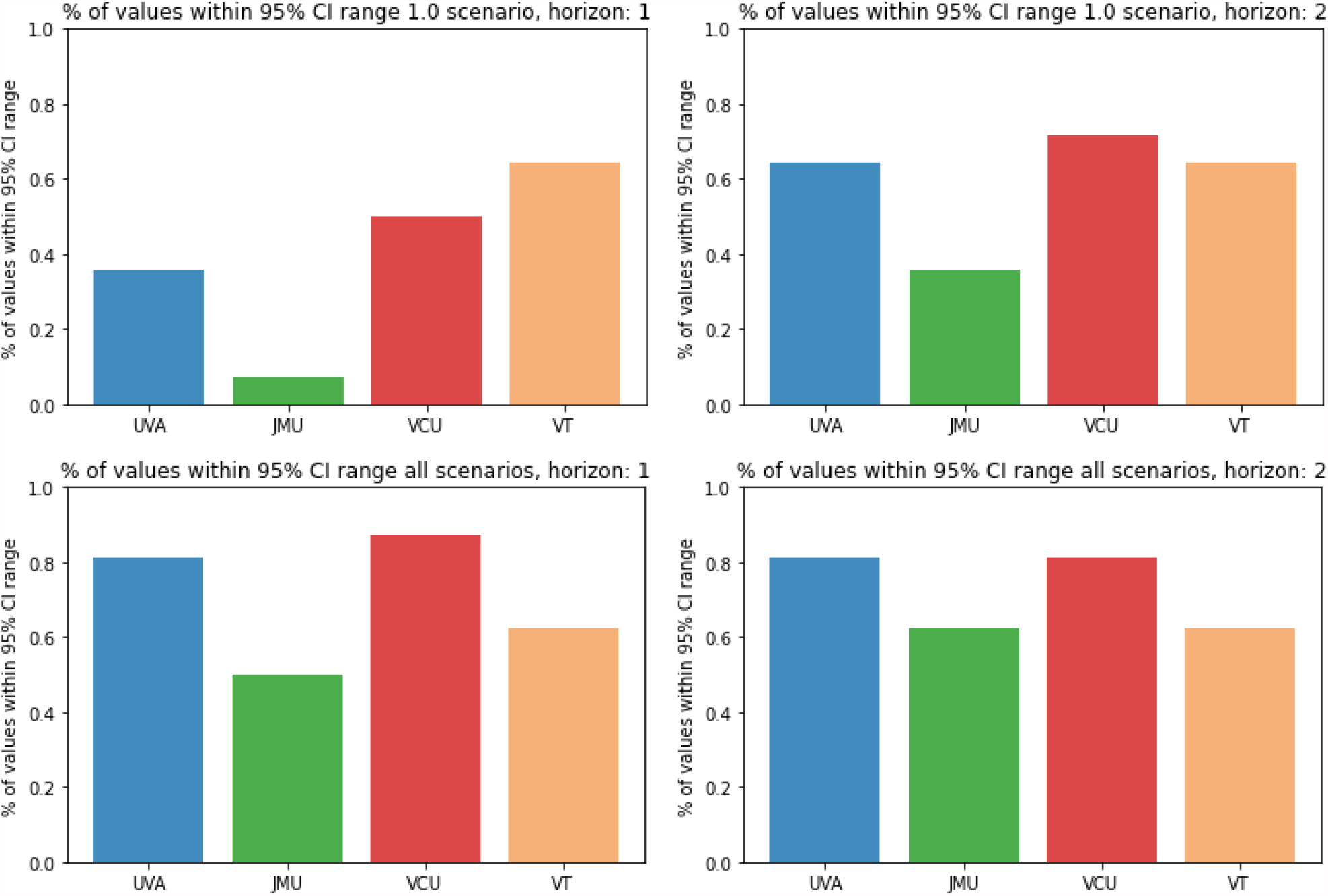
The above shows percent of predictions whose 95% CI band contain the realized cases for the one week and two week horizons. The 1.0 scenario only, shown on top. The capture from any of the three scenarios shown below.

The case forecasts inform the process that regulates interactions, but in particular can help the college manage the number of students in quarantine, and the demands on those facilities. If the college has the ability to adjust its isolation or quarantine capacity, an increasing or decreasing case count and model forecasts indicating a worsening or easing of the new case trend is useful information for the college in deciding to whether to change the capacity of their facilities. In addition, the isocurves allow the school to manage the frequency of testing the student population. Combined, the school is able to find the optimal testing frequency and management of other intervention measures such that the forward case flow is tenable to the administration, when considering isolation facilities, etc.

When considering the application of this work going forward, of primary interest is adding more schools to the analysis and expanding the scope to the national level. Including schools from across the country (which will help account for regional differences in COVID responses) and across various sizes of schools (which will help account for resource differences in implementing interventions, communication, and student accommodations) will provide more information on the usefulness of the model for all schools. The SEIR model and an updated technical report can be found at https://github.com/benjaminhurt/University_SEIR_Model.

## Data Availability

Data and code will be made available upon request.

## Acknowledgements

The authors would like to thank members of the Network Systems Science and Advanced Computing (NSSAC) Division for their thoughtful comments and suggestions related to epidemic modeling and response support. We thank members of the Biocomplexity Institute and Initiative, University of Virginia, for useful discussion and suggestions. This work was partially supported by National Institutes of Health (NIH) Grant 1R01GM109718, NSF BIG DATA Grant IIS-1633028, NSF Grant No.: OAC-1916805, NSF Expeditions in Computing Grant CCF-1918656, CCF-1917819, NSF RAPID CNS-2028004, NSF RAPID OAC-2027541, US Centers for Disease Control and Prevention 75D30119C05935, a grant from Google, University of Virginia Strategic Investment Fund award number SIF160, Defense Threat Reduction Agency (DTRA) under Contract No. HDTRA1-19-D-0007, and Virginia Dept of Health Grant VDH-21-501-0141.

**BENJAMIN HURT** is a Data Scientist at Biocomplexity. He earned his Masters in Data Analytics from the Georgia Institute of Technology. His research interests include machine learning, artificial intelligence, forecasting, and financial system analysis. At NSSAC, his focus has been the development of epidemiological forecasting and analysis, along with financial contagion modeling. His email address is.

**ANIRUDDHA ADIGA** is a Postdoctoral Research Associate at the NSSAC Division of the Biocomplexity Institute and initiative. He completed his Ph.D. from the Department of Electrical Engineering, Indian Institute of Science (IISc), Bangalore, India and has held the position of Postdoctoral fellow at IISc and North Carolina State University, Raleigh, USA. His research areas include signal processing, machine learning, data mining, forecasting, big data analysis, etc. At NSSAC, his primary focus has been the analysis and development of forecasting systems for epidemiological signals such as influenza-like illness and COVID-19 using auxiliary data sources. His email address is.

**MADHAV MARATHE** is a Distinguished Professor in Biocomplexity, the division director of the Networks, Simulation Science and Advanced Computing Division at the Biocomplexity Institute and Initiative, and a Professor in the Department of Computer Science at the University of Virginia (UVA). His research interests are in network science, computational epidemiology, AI, foundations of computing, socially coupled system science and high performance computing. Before joining UVA, he held positions at Virginia Tech and the Los Alamos National Laboratory. He is a Fellow of the IEEE, ACM, SIAM and AAAS. His email address is.

**CHRISTOPHER L. BARRETT** is an endowed Distinguished Professor in Biocomplexity, the Executive Director of the Biocomplexity Institute, and Professor of the Department of Computer Science at the University of Virginia. He is an interdisciplinary computational scientist who has published more than 100 research articles exploring all aspects of large multiscale interaction systems. Over the past 35 years, Barrett has conceived, founded and led large interdisciplinary complex systems research projects and organizations, established national and international technology programs, and co-founded organizations for federal agencies such as the Department of Defense, the Department of Energy and the Department of Homeland Security. He has served in various advisory and collaborative scientific roles internationally. His email address is.

